# Community-based causal evidence that high habitual caffeine consumption alters distinct polysomnography-derived sleep variables

**DOI:** 10.1101/2024.12.11.24318776

**Authors:** Benjamin Stucky, Leonard Henckel, Marloes H. Maathuis, José Haba-Rubio, Pedro Marques-Vidal, Francesca Siclari, Raphaël Heinzer, Hans-Peter Landolt

## Abstract

Caffeine is the most widely consumed psychoactive substance in the World and acutely impairs sleep quality when studied in the laboratory. The repercussions on sleep of chronic caffeine intake, which is common in society, are unknown. Because adequate sleep is important for general health, we explored causal effects of habitual caffeine intake on objective and subjective sleep quality with two-sample Mendelian Randomization and matching methods. We analyzed large community-based datasets taken from the UK Biobank (n=485,511) and HypnoLaus (n=1,702) cohorts. We consistently show that intake of four or more caffeinated beverages daily shortens total sleep time compared to three or fewer caffeinated drinks per day. While we found shorter sleep with all models, the estimated reductions varied from 11-229 minutes, likely due to modest instrument strengths. A concurrent increase in sleep intensity indicates at least partial compensation. The findings suggest that high daily caffeine intake causally alters sleep patterns in the general population.

## Introduction

Caffeine use in humans has a rich and long history^1^. Today, caffeine is considered the most widely consumed psychoactive substance in the world^2^. It occurs naturally and is readily available in common foods and beverages, including coffee, tea, energy drinks and chocolate^3^. Consumers often do not know the caffeine content of their nutrition^4,5^. Based on estimates from wastewater analyses, the average habitual daily caffeine consumption in Europe ranges from 86-263 mg per person, which equals roughly 1-3 cups of coffee^6^. Motivations for caffeine intake include the substance’s promotion of alertness, taste and smell, convivial social settings, increased physical energy, and alleviation or management of stress^7,8^. Interestingly, genetic factors modulate some individual differences in caffeine effects and consumption^9–12^.

The prevalent use of caffeine raises questions regarding its potential health consequences. A majority of studies concluded that there are no major adverse health effects of moderate caffeine intake in the adult general population. On the contrary, caffeine consumed in normal doses was associated with reduced risk for several chronic diseases^13–18^. Unfortunately, the available evidence often relies on observational and prospective studies that are not well suited to derive causality.

A particularly interesting aspect of caffeine-related health is the quality of sleep^19^. Undisturbed sleep is required for many physical and cerebral processes, including cardiovascular, immune and memory processes, as well as other functions like emotion regulation^20–24^. Carefully controlled polysomnographic recordings including electroencephalographic (EEG) measurements revealed that acute caffeine intake prolongs the time to fall sleep and wakefulness after sleep onset, and reduces sleep efficiency, sleep duration and non-rapid-eye-movement (NREM) sleep intensity^25–27^. However, such studies typically enroll habitual caffeine consumers and instruct them to abstain prior to the experiment. Hence, they investigate the effects of caffeine after abstinence, which may unmask possible adaptations to the chronic intake^28^, which is common in society^29^.

Very little is currently known about the effects of chronic caffeine intake on sleep. Intriguingly, some evidence suggests that repeated caffeine intake increases rather than decreases NREM sleep intensity. Specifically, a study in mice revealed that two weeks of continuous caffeine consumption enhanced both the amplitude of the daily sleep-wake cycle as well as behavioral and EEG markers of sleep intensity^30^. Similarly, in healthy men, repeated chronic caffeine intake over ten consecutive days did not reduce EEG-derived NREM sleep intensity but delayed the occurrence of REM sleep^28,31^. Taken together, the data collected in laboratory settings may suggest that chronic caffeine consumption induces tolerance to the acute effects on sleep and the sleep EEG mentioned above^32–35^. Nonetheless, controlled laboratory protocols have limitations to elucidate habitual caffeine effects on sleep because they cannot take into account the large variation in population habits that often persist for years. In addition, home sleep recordings have the advantage that the participants sleep in their familiar environment rather than in an unknown and artificial laboratory setting and can maintain their regular routines.

A promising approach to address these limitations in assessing causal effects is the use of statistical tools such as Mendelian Randomization (MR) in epidemiological datasets^36–39^. These methods employ genetic variants associated with an exposure variable as instruments, to estimate causal effects on an outcome variable. The exposure variable in our study is habitual caffeine intake, which is modulated by genetic influences^9–11,40,41^ and lends itself to MR methodologies. The main idea behind MR is that genetic variants are to a certain degree randomly inherited and stable throughout life. Hence, they can be seen as part of a natural experiment that mimics a randomized controlled trial. Previous MR research on the relationships between habitual caffeine intake and sleep-associated outcomes^42,43^ relied on subjective sleep estimates, which can be systematically biased and often deviate from objective measures^44–46^.

By contrast, causal effects of habitual caffeine intake on objective markers of sleep quality are unknown. To start filling this gap of knowledge, we estimated the causal effects of habitual caffeine consumption on objective and subjective sleep variables recorded at home, using three distinct two-sample MR methods in the two large cohorts UK Biobank^47^ and HypnoLaus^46,47^.

## Results

First, we estimated the associations of relevant single nucleotide polymorphisms (SNPs) with the amount of habitual caffeine intake in the UK Biobank dataset^47^. We based this analysis on the subset of data that provide both genetic information as well as self-reported caffeine intake (n = 485’511). Next, we explored the HypnoLaus dataset^46,47^ (n = 1’755) for associations between the same SNPs and sleep recorded at the participants’ homes. The demographic characteristics of the study participants are summarized in Table 1 (see also supplementary Table S1). The distributions of self-reported, habitual caffeine intake in the UK Biobank and HypnoLaus cohorts are comparable, with a majority of participants reporting habitual intake of 1-3 caffeinated beverages per day (Fig. 1).

**Table 1.**
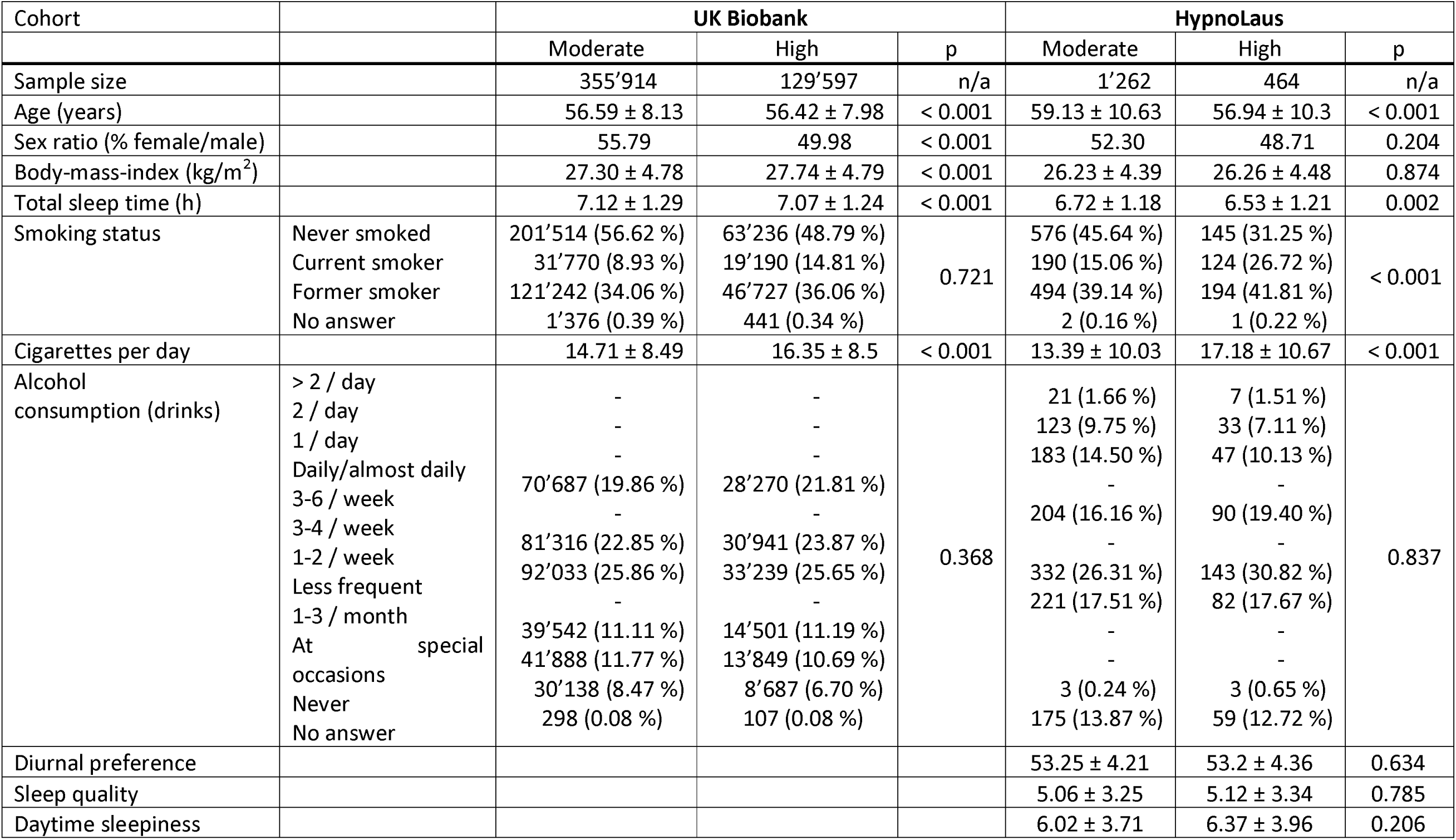
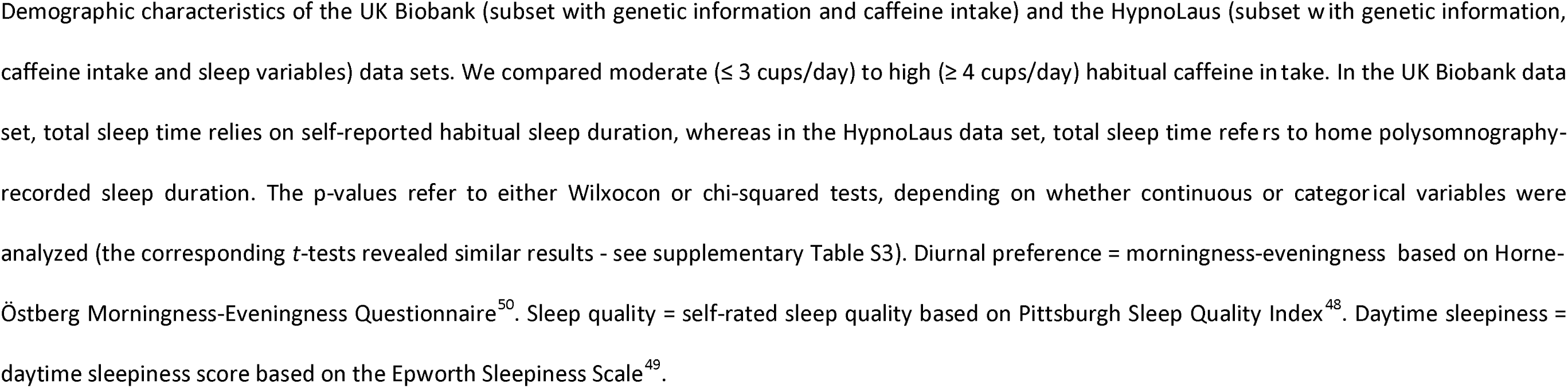
Demographics.

| Cohort |  | UK Biobank |  |  | HypnoLaus |  |  |
| --- | --- | --- | --- | --- | --- | --- | --- |
|  |  | Moderate | High | p | Moderate | High | p |
| Sample size |  | 355'914 | 129'597 | n/a | 1'262 | 464 | n/a |
| Age (years) |  | 56.59 ± 8.13 | 56.42 ± 7.98 | < 0.001 | 59.13 ± 10.63 | 56.94 ± 10.3 | < 0.001 |
| Sex ratio (% female/male) |  | 55.79 | 49.98 | < 0.001 | 52.30 | 48.71 | 0.204 |
| Body-mass-index (kg/m<sup>2</sup>) |  | 27.30 ± 4.78 | 27.74 ± 4.79 | < 0.001 | 26.23 ± 4.39 | 26.26 ± 4.48 | 0.874 |
| Total sleep time (h) |  | 7.12 ± 1.29 | 7.07 ± 1.24 | < 0.001 | 6.72 ± 1.18 | 6.53 ± 1.21 | 0.002 |
| Smoking status | Never smoked<br>Current smoker<br>Former smoker<br>No answer | 201'514 (56.62 %)<br>31'770 (8.93 %)<br>121'242 (34.06 %)<br>1'376 (0.39 %) | 63'236 (48.79 %)<br>19'190 (14.81 %)<br>46'727 (36.06 %)<br>441 (0.34 %) | 0.721 | 576 (45.64 %)<br>190 (15.06 %)<br>494 (39.14 %)<br>2 (0.16 %) | 145 (31.25 %)<br>124 (26.72 %)<br>194 (41.81 %)<br>1 (0.22 %) | < 0.001 |
| Cigarettes per day |  | 14.71 ± 8.49 | 16.35 ± 8.5 | < 0.001 | 13.39 ± 10.03 | 17.18 ± 10.67 | < 0.001 |
| Alcohol consumption (drinks) | > 2 / day<br>2 / day<br>1 / day<br>Daily/almost daily<br>3-6 / week<br>3-4 / week<br>1-2 / week<br>Less frequent<br>1-3 / month<br>At special occasions<br>Never<br>No answer | -<br>-<br>-<br>70'687 (19.86 %)<br>-<br>81'316 (22.85 %)<br>92'033 (25.86 %)<br>-<br>39'542 (11.11 %)<br>41'888 (11.77 %)<br>30'138 (8.47 %)<br>298 (0.08 %) | -<br>-<br>-<br>28'270 (21.81 %)<br>-<br>30'941 (23.87 %)<br>33'239 (25.65 %)<br>-<br>14'501 (11.19 %)<br>13'849 (10.69 %)<br>8'687 (6.70 %)<br>107 (0.08 %) | 0.368 | 21 (1.66 %)<br>123 (9.75 %)<br>183 (14.50 %)<br>-<br>204 (16.16 %)<br>-<br>332 (26.31 %)<br>221 (17.51 %)<br>-<br>-<br>3 (0.24 %)<br>175 (13.87 %) | 7 (1.51 %)<br>33 (7.11 %)<br>47 (10.13 %)<br>-<br>90 (19.40 %)<br>-<br>143 (30.82 %)<br>82 (17.67 %)<br>-<br>-<br>3 (0.65 %)<br>59 (12.72 %) | 0.837 |
| Diurnal preference |  |  |  |  | 53.25 ± 4.21 | 53.2 ± 4.36 | 0.634 |
| Sleep quality |  |  |  |  | 5.06 ± 3.25 | 5.12 ± 3.34 | 0.785 |
| Daytime sleepiness |  |  |  |  | 6.02 ± 3.71 | 6.37 ± 3.96 | 0.206 |

**Figure 1.**
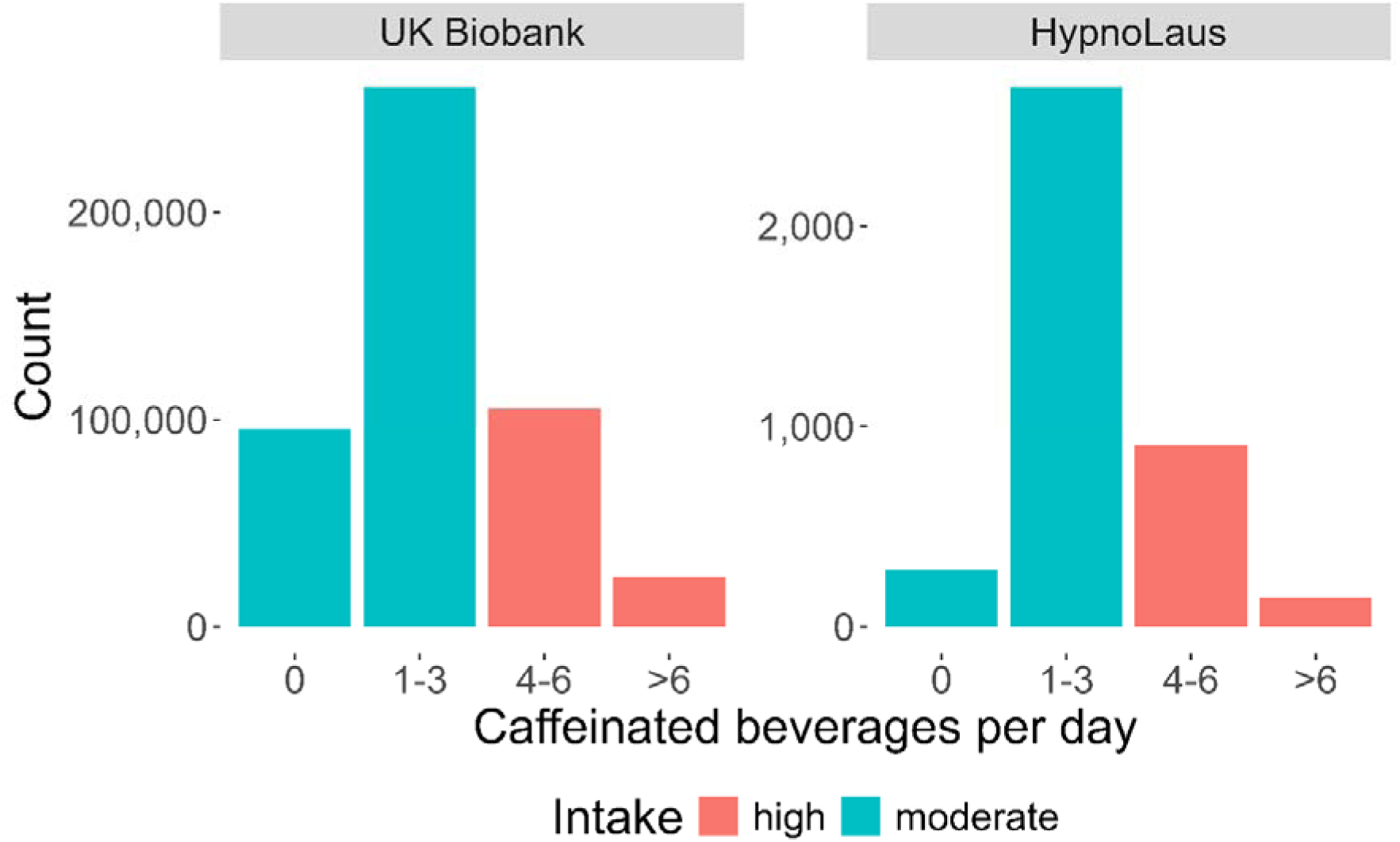
Distributions of self-reported intake of caffeinated beverages per day in participants of the UK Biobank (n = 485,511) and HypnoLaus (n = 1,702) cohorts. The color code highlights the high (red; ≥ 4 caffeinated beverages per day) and moderate (blue; ≤ 3 caffeinated beverages per day) intake groups. Please note the different scales (y-axes) between the two data sets.

To comprehensively capture the multifaceted aspects of sleep quality, we analyzed the following objective variables: total sleep time, sleep latency (i.e., time from lights-out to first occurrence of NREM sleep stage N2), number of awakenings, percentage of rapid-eye-movement (REM) sleep, and the proportion of delta (1-4 Hz) and sigma (12-16 Hz) power in the NREM sleep EEG. As subjective measures, we analyzed the amply validated Pittsburgh Sleep Quality Index (PSQI)^48^, the Epworth Sleepiness Scale (EES)^49^ of daytime sleepiness and the Morningness-Eveningness-Questionnaire (MEQ)^50^ to estimate diurnal preference. The distributions of all outcome variables can be seen in Fig. 2. They show high variability in objective and subjective sleep quality measures as expected in a heterogenous, community-derived study sample.

**Figure 2.**
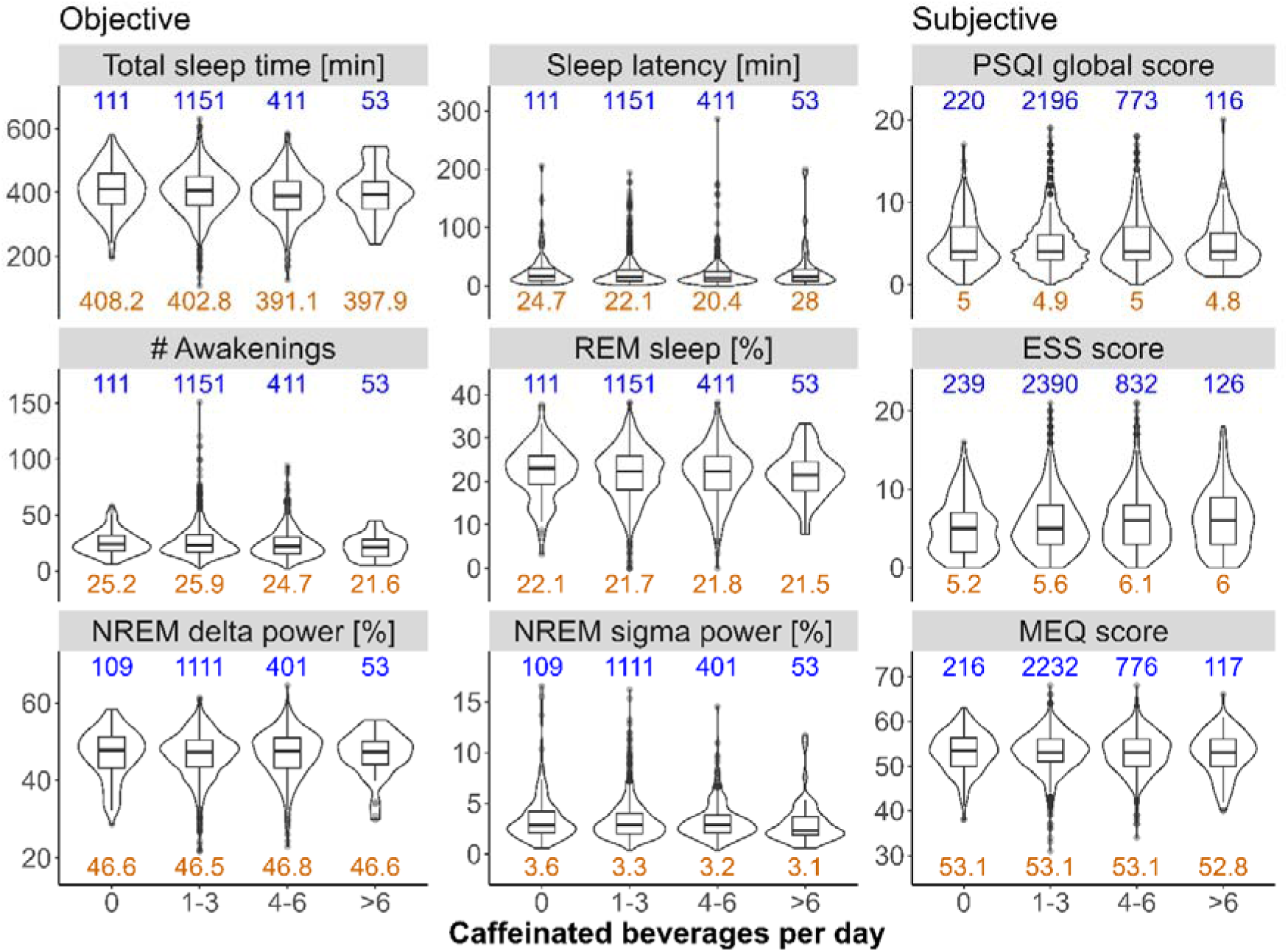
Distributions of sleep variables collected in the HypnoLaus data set (n = 1,702). The left and middle panels illustrate home polysomnography-derived, objective sleep quality: total sleep time (min), sleep latency (min, time between lights-out and first occurrence of stage N2 sleep), number (#) of awakenings, REM sleep (expressed as % of total sleep time), EEG delta power in NREM sleep (spectral power in the 1-4 Hz range expressed as a % of total power), and EEG sigma power in NREM sleep (spectral power in the 12-16 Hz range expressed as a % of total power). The right panel illustrates self-reported measures of sleep quality: Pittsburgh Sleep Quality Index (PSQI, global score), Epworth Sleepiness Scale (ESS) score, and Morningness-Eveningness Questionnaire (MEQ) score. X-axes: self-reported intake of caffeinated beverages per day. The blue values on top of each panel indicate the sample size per group. The orange values on the bottom of each panel indicate the mean value of the corresponding distribution.

### Mendelian Randomization

We estimated causal differences in sleep quality between the high (self-reported ≥ 4 caffeinated beverages per day) and moderate (≤ 3 caffeinated beverages) caffeine intake groups. Because each MR method requires a distinct set of assumptions that are difficult to verify, we employed three different, complementary methods: MR-Egger^51^, inverse variance weighting (IVW)^38^, and weighted median^52^. All three methods revealed no differences between the groups in sleep latency, the percentage of REM sleep, sigma power in NREM sleep, nor self-rated sleep quality (PSQI global score) (Fig. 3). One or two, but not all three MR methods, suggested an effect of high caffeine intake on the number of awakenings (MR-Egger: estimate = 1.102, p = 0.038; weighted median: estimate = 0.64, p = 0.01), delta power in NREM sleep (IVW: estimate = 8.82%, p = 0.02; weighted median: estimate = 10.25%, p = 0.002), daytime sleepiness (ESS score) (IVW: estimate = −0.85, p = 0.04), and diurnal preference (MEQ) (weighted median: estimate = 8.34, p = 0.003). When looking at total sleep time, all three methods showed shorter sleep in individuals reporting the consumption of more than three caffeinated beverages per day (MR-Egger: estimate = −229 min, p = 0.03; IVW: estimate = −125 min, p = 0.005; weighted median: estimate = −140 min, p < 0.001) (Fig. 3).

**Figure 3.**
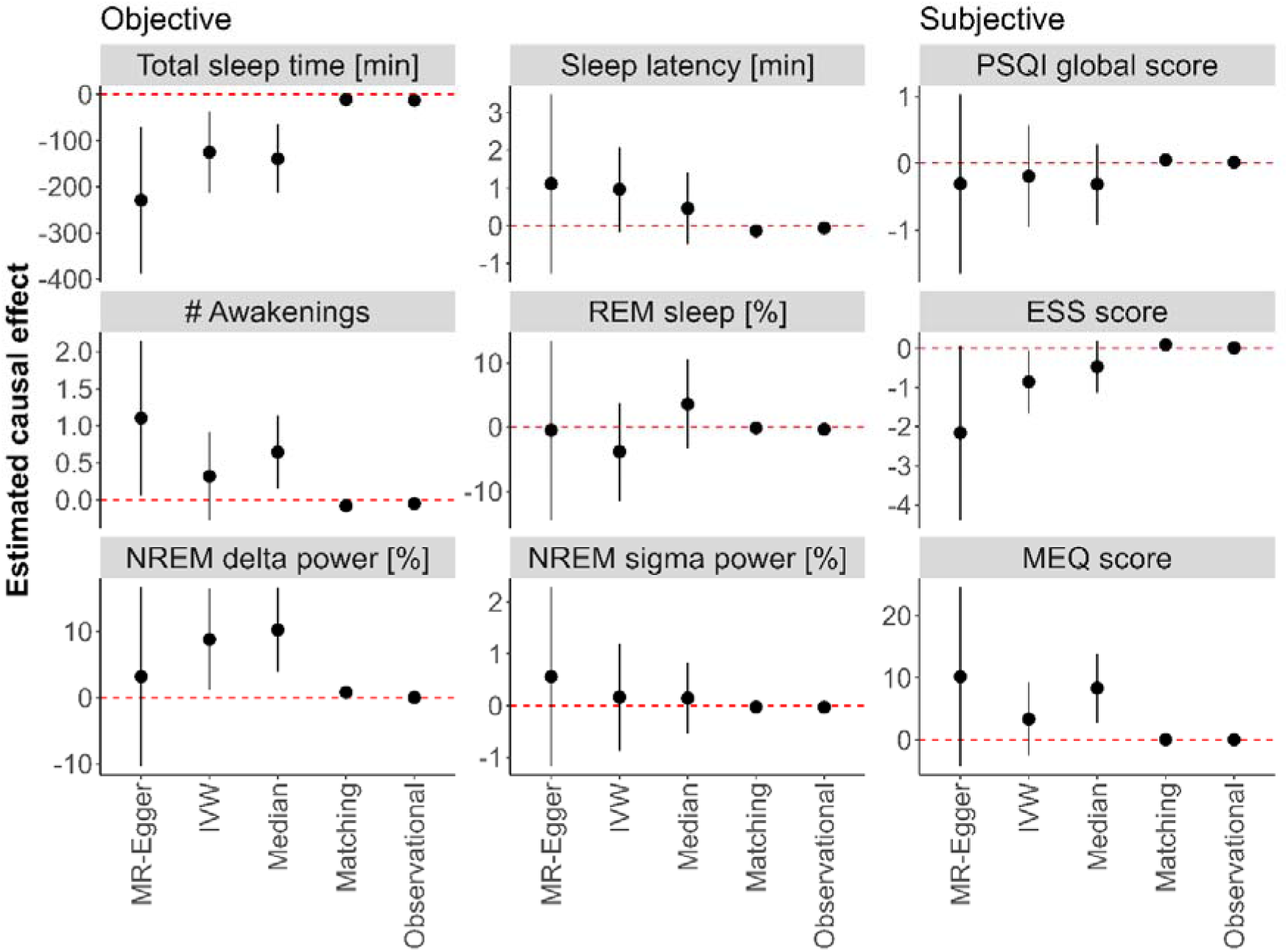
Mendelian randomization (MR), causal matching and observational estimators between participants in the HypnoLaus data set (n = 1,702) reporting high (black dots; ≥ 4 caffeinated beverages per day) and moderate (red vertical dashed lines at 0; ≤ 3 caffeinated beverages per day) habitual caffeine intake. The left and middle panels illustrate home polysomnography-derived, objective sleep quality: total sleep time (min), sleep latency (min, time between lights-out and first occurrence of stage N2 sleep), number (#) of awakenings, REM sleep (expressed as % of total sleep time), EEG delta power in NREM sleep (spectral power in the 1-4 Hz range expressed as a % of total power), and EEG sigma power in NREM sleep (spectral power in the 12-16 Hz range expressed as a % of total power). The right panel illustrates self-reported measures of sleep quality: Pittsburgh Sleep Quality Index (PSQI, global score), Epworth Sleepiness Scale (ESS) score, and Morningness-Eveningness Questionnaire (MEQ) score. Estimators include the MR methods MR-Egger, inverse variance weighting (IVW) and weighted median (Median), the MatchingFrontier algorithm (Matching) and the observational effect (Observational). Error bars refer to the 95% confidence intervals. The values on the y-axes were log-transformed for sleep latency, number of awakenings, PSQI global score, and ESS score. The values of NREM sigma power are logit-transformed. Other values were not transformed.

After correction for multiple testing with the Holm-Bonferroni adjustment^53^, the weighted median estimate for delta power in NREM sleep (p = 0.044) and total sleep time (p = 0.006) remained significant. All three MR-estimators exhibited high uncertainty, likely due to the inherently weak genetic instruments. This was particularly true for the MR-Egger estimator. Thus, the high mean difference (> two hours) between the high and moderate caffeine consumers estimated with these methods appears unrealistically large.

### Secondary Analyses

To corroborate the MR results and to estimate a realistic effect size between high and moderate chronic caffeine intake, we also employed the MatchingFrontier method for causal matching^54^. To place the causal estimates into context, we also computed the observational effects. The causal matching revealed no differences for the percentage of REM sleep, sigma power in the NREM sleep EEG, subjective sleep quality, and diurnal preference. By contrast, it showed shorter sleep latency (estimate = −0.1 min p = 0.003), lower number of awakenings (estimate = −0.079, p = 0.003), and enhanced daytime sleepiness (estimate = 0.097, p < 0.001) in the high caffeine intake group. Corroborating the results of the MR, causal matching confirmed a shorter total sleep time per night (estimate = −11 min, p = 0.007) and higher delta power in the NREM sleep EEG (estimate = 0.8%, p = 0.018) in the high caffeine intake group. Although the effect sizes were rather small (total sleep time: Cohen’s *d* = −0.15; delta power: Cohen’s *d* = 0.14), the difference in total sleep time was strikingly similar to the observational effect (12.9 min, p < 0.001; Fig. 3).

For the other variables analyzed, the observational effects did not suggest a significant difference between the groups. We report the complete results, including estimates, standard errors, confidence intervals, p-values and Cohen’s d coefficients in supplementary Table S2.

## Discussion

To elucidate the causal impact of consuming ≥ 4 caffeinated beverages per day on complementary aspects of objective and subjective sleep quality, we applied three distinct MR methods, as well as causal matching and observational effects, in two large datasets of community-based cohorts in the UK and Switzerland. We found mild but consistent evidence for causally reduced total sleep time in individuals with high habitual caffeine intake when compared to individuals with a moderate habitual consumption of ≤ 3 caffeinated beverages per day. While the difference between the groups was probably overestimated with MR, likely due to moderate associations of the genetic instruments, the causal matching and observational estimates suggest a shortening of sleep by roughly 11-13 min per night. Given the importance of adequate sleep for many important health outcomes^19^, even this seemingly small difference in total sleep time warrants consideration^55^.

Previous randomized studies found no significant effects of consuming two to three doses of caffeine daily on total sleep time^28,31,56^. However, the previous work investigated short durations of repeated caffeine administration (i.e., 9 and 14 days) in small samples of 11 and 20 participants. By design, they either administered caffeine no later than lunch, scheduled strict bedtimes, or measured sleep eight hours after last caffeine ingestion^28,31,56^. These laboratory settings do not accurately reflect the natural sleep patterns and habitual caffeine intake of the general population, and are unable to assess effects that only become apparent after long-term consumption^57^.

Another trial employed a randomized, cross-over, yet open design, and estimated sleep duration across 14 days with a wrist-worn, commercial fitness tracker in 100 adults. The participants were randomly assigned for consecutive 2-day periods to either drink coffee or not^58^. Estimated sleep duration was 36 min shorter on caffeine days when compared to non-caffeine days. Although this estimate roughly corresponds to our finding, intermittent caffeine intake and abstinence cannot exclude carry-over and withdrawal effects. Furthermore, wearable-derived estimates of sleep may be systematically biased^59^ and have low accuracy in individual recordings^60^.

A previous two-sample MR study on the effects of caffeine consumption on self-reported sleep variables was rather inconclusive^43^. The authors sampled sleep duration with a poor temporal resolution (in hours), which weakens causal estimates. In addition, self-reports are prone to subjective-objective sleep discrepancy and differ among diverse study populations^61,62^. In accordance with our study, they did not find a causal effect of caffeine on subjective sleep quality. Together, the two studies indicate that objective and subjective measures may capture distinct aspects of sleep quality.

Previous controlled experiments in rodents and humans revealed EEG-derived evidence that repeated caffeine intake over several days can enhance sleep pressure when compared to placebo^28,30,31,63^. In accordance with these findings, two different MR methods (IVW and median weighted) suggested higher delta power in NREM sleep in the high caffeine group when compared to the moderate caffeine group. The median weighted method also withstood correction for multiple testing. Although the difference between the groups pointed in the same direction, the MR-Egger estimate was not significant. Nonetheless, this result was confirmed by the causal matching analysis for delta power in NREM sleep. Together, the findings raise the intriguing possibility that chronically reduced total sleep time caused by high caffeine consumption may be compensated by slightly enhanced sleep intensity as measured in the sleep EEG. This regulatory principle is well known from controlled experiments^64^. Its confirmation in large community-based studies would suggest that habitual caffeine use does not impact the homeostatic aspect of sleep-wake regulation, which is consistent with the notion that caffeine cannot compensate for lost sleep. It would be interesting to corroborate this notion with polysomnographic recordings in large and diverse participant samples, yet such measurements are time consuming, highly expensive, and not readily available. Easier available wearable sleep recordings in large genetic cohorts cannot unequivocally clarify this question.

It needs to be mentioned that several limitations apply to this work. First, the treatment variable (cups of caffeinated beverages per day) was self-reported and, thus, susceptible to inaccurate recall or reporting.^65^ Additionally, we averaged over beverages with different caffeine contents and other dietary sources of caffeine may have been missed.^66^ The large sample sizes weaken this concern. Thus, the distributions of reported caffeine use in the two cohorts correspond to previous studies^57^ and quantitative wastewater measurements^6^. Therefore, we expect that no systematic reporting bias was present. Second, instrumental variable estimators typically suffer from low accuracy, especially if the instruments are weak. Weak instruments reduce statistical power to detect causal effects. This concern not only applies to the present study, but to MR studies in general. By contrast, causal matching does not suffer from weak instruments but from potential unobserved confounding. Thus, we complemented different MR methods with causal matching and found convergent evidence for reduced total sleep time and increased delta power caused by high habitual caffeine intake. Third, the assumptions of different MR methods are often difficult, if not impossible, to verify. For example, we selected the instruments based on current expert knowledge of the gene variants contributing to differences in caffeine intake and sleep quality. Nevertheless, it remains often uncertain whether an instrument is valid, because gene variants can affect the outcome in another way than only via the treatment. This phenomenon is referred to as pleiotropy. The intercepts in the MR-Egger regression analysis showed no evidence for pleiotropy. In addition, we employed three distinct MR methods and a causal matching estimator, to reduce the risk that violated MR assumptions skew the results. Fourth, most participants in the two cohorts are of European descent. Large studies in more diverse populations are necessary to generalize the present results. Finally, we don’t know whether a reduced propensity to nap in the high caffeine intake group could have contributed to the observed increase in NREM delta power because naps are not recorded in the HypnoLaus cohort.

In conclusion, we used four complementary statistical methods in two large, high-quality, community-based data sets, to estimate the causal effects of chronic high caffeine intake on objective and subjective sleep quality estimates recorded at home. Compared to people consuming three or less caffeine containing beverages per day, we found consistent evidence for reduced total sleep time and increased sleep intensity in people who habitually consume more than four caffeinated drinks per day. Although the differences between high and moderate caffeine users appear small, these novel insights are relevant and warrant further investigations because habitual daily caffeine intake is a ubiquitous human behavior.

## Methods

### Study population

This study was conducted using the UK Biobank and HypnoLaus cohorts. The UK Biobank is a very large open access prospective study conducted in the UK^47^. We used the subpopulation that contains information on caffeine intake and genetic information, amounting to a total of 485’511 participants. The UK Biobank was used to estimate the association of various gene variants with caffeine intake. The selection process is described below. The HypnoLaus cohort, which belongs to the CoLaus/ PsyCoLaus cohort, is a population-based study of Lausanne, Switzerland^67,68^. The HypnoLaus data set contains caffeine intake, genetic and objective sleep data collected from at-home polysomnography in 1’755 participants. The HypnoLaus cohort was used to estimate the association of each gene variant with several objective and subjective measure of sleep quality.

### Caffeinated beverages per day

The variable caffeinated beverages per day was measured in the HypnoLaus data-set with granularity: 0 cups/day, 1 – 3 cups/day, 4-6 cups/day, >6 cups/day. The UK Biobank contained more detailed information on caffeine intake at a higher granularity, for example different beverage types (tea, coffee, caffeinated sodas, etc) were listed. For comparability, we combined the intake of these various beverages types and reduced the intake detail to match the cups of caffeinated beverage intake categories from the HypnoLaus data-set. The resulting distributions are shown in Fig. 1. To streamline the MR analyses and obtain more balanced caffeine intake groups, we split the intake habits into high (≥ 4 caffeinated drinks/day) and moderate (≤ 3 caffeinated drinks/day) caffeine use.

### Sleep variables in the HypnoLaus dataset

Participants performed a full night PSG at home (Titanium, Embla® Flaga, Reykjavik, Iceland). PSG were performed according to the American Academy of Sleep Medicine (AASM) 2007 recommendations ^69^ and included: electroencephalography (EEG) leads (F3, F4, C1, C2, O1 and O2, 256 Hz sampling rate); electrooculography (EOG, left and right); electromyography (EMG, chin and anterior tibialis muscle); electrocardiography (ECG, one lead); oxygen saturation (SpO_2_); airflow (nasal cannula); abdominal and thoracic respiratory efforts; snoring; and body position. PSG data were visually scored according to the AASM guidelines 2007 ^69^ for sleep.

We considered the following six objective measure of sleep quality: Total sleep time, sleep latency (time between lights-off to N2 sleep), number of awakenings, percentage of REM sleep per total sleep time, and EEG delta (1-4 Hz) and sigma (12-16 Hz) power in NREM sleep relative to the total power between 0.5 and 30 Hz. We prospectively selected these distinct sleep measures, to objectively capture different aspects of the complex and multifaceted quality of sleep. For a detailed explanation of the EEG data preprocessing and artifact handling, please see refer to reference^70^.

To complement our analyses and verify previous studies relying on subjective sleep measures^43^, we also included the following validated sleep quality questionnaires: Pittsburgh Sleep Quality Index (PSQI, global score)^48^, Epworth Sleepiness Scale (ESS)^49^, and Morningness-Eveningness Questionnaire (MEQ)^50^. The PSQI is the most commonly used self-perceived sleep quality questionnaire, the ESS is the most common subjective instrument to capture daytime sleepiness, and the MEQ is used to assess the individual’s preference for being active in the morning or evening, also referred to as diurnal preference.

### Mendelian Randomization (MR)

Instrumental variable estimators can estimate causal effects of a treatment on an outcome even in the presence of unmeasured confounding or reverse causality, i.e., the outcome causing the treatment^71,72^. In general, they require an auxiliary variable, referred to as instrument, which has to i) be robustly associated with the treatment; ii) not share common causes with the outcome; and iii) affect the outcome only via the treatment. While precondition i) can be verified statistically, ensuring preconditions ii) and iii) requires subject matter knowledge^38,73^. Mendelian Randomization is a special case of an instrumental variable estimator, where a gene variant is used as an instrument^38^. Gene variants (i.e., single nucleotide polymorphism, SNPs) are natural instruments because they cannot be affected by most observational covariates. As a result, precondition ii) holds for most potential treatments and outcomes. Precondition iii), which is often referred to as the no-pleiotropy assumption, can still be violated. Thus, the SNPs should be carefully selected to minimize the risk of violating this assumption. To further protect against violations of the no-pleiotropy assumption, newer MR methods typically use multiple instruments^51,52,74^. We used three such estimators. First, the MR-Egger estimator, which is consistent under the InSIDE (instrument strength independent of direct effect) assumption. That is, the association between the outcome and the instruments is independent of the association between the treatment and the instruments. In principle, InSIDE may hold even if all SNPs are invalid. Second, the inverse variance weighted (IVW) average of Wald ratios^38^. This estimator is consistent if all individual SNPs are valid. The IVW average has the major advantage that it is more statistically accurate than any of the individual Wald ratios, which tend to suffer from low accuracy^75^. And third, the weighted median of Wald ratios^52^. This estimator is consistent if more than half of the weight comes from valid instruments. Thus, the weighted median is robust against some invalid instruments.

We used a two-sample MR approach. This procedure can have some advantages compared to one-sample MR. It can increase statistical power to detect effects, reduce bias due to weak instruments, and prevent underestimation of causal effects due to the winner’s curse^76^. Two-sample MR methods assume that the two study populations are comparable.^76^

### Causal Matching

To complement the MR methods, we also employed causal matching to estimate the causal effects of high habitual caffeine consumption on objective and subjective measures of sleep quality. A randomized controlled trial between two groups of interest (here, high and moderate caffeine intake groups) seeks to ensure that all other observed and unobserved covariates are equally distributed in the groups. This approach is the ideal setting to assess causal effects. In an observational setting, however, people with high habitual caffeine intake likely also differ in other aspects from people with moderate caffeine intake. A possible solution to this problem is to match each participant in the high intake group to a participant in the low intake group with similar characteristics, to achieve a “pseudo” randomization^77^. However, this approach may fail if some common causes of the treatment and outcome have not been measured. Because this likely is the case here, e.g., due to unmeasured interindividual differences in circadian rhythms or lifestyle factors, we used matching merely to corroborate the MR results.

We used MatchingFrontier^54^, which simultaneously optimizes the similarity between the groups of interest and the matched sample size. To obtain good balance, we only retained 50% of the best matches. The exact percentage of retained matches did not strongly affect the results (supplementary Fig. S1). Depending on the outcome variable, when retaining 50% of the matches, 385-792 observations per variable remained.

We used various covariates available in the data sets for matching. They included physical health (subjective overall health rating, total weekly energy expenditure excluding sleep, alcohol units per week, smoking cigarettes equivalent per day, body mass index); socio-demographic status (age, gender, self-reported ethnicity, highest education level, marital status); as well as economic characteristics (occupational position).

### Single nucleotide polymorphism selection

Based on a literature search of prior publications investigating the association of genetic variants on caffeine consumption, we preselected 83 SNPs. The references and selected SNPs can be found in supplementary Table S3. In the UK Biobank data, we calculated the t-value of each SNP association with caffeine intake. Due to missing SNP information, we could not calculate the t-values for 6 of the preselected SNPs. To mitigate power loss due to weak instruments, an absolute t-value > 8 was used as inclusion criterion for the MR analyses. This value corresponds to a slightly stricter cut-off value as the common 5 × 10^−8^ genome-wide significance p-value threshold^78^. This resulted in 30 SNPs selected for the analyses (Table 2).

**Table 2.**
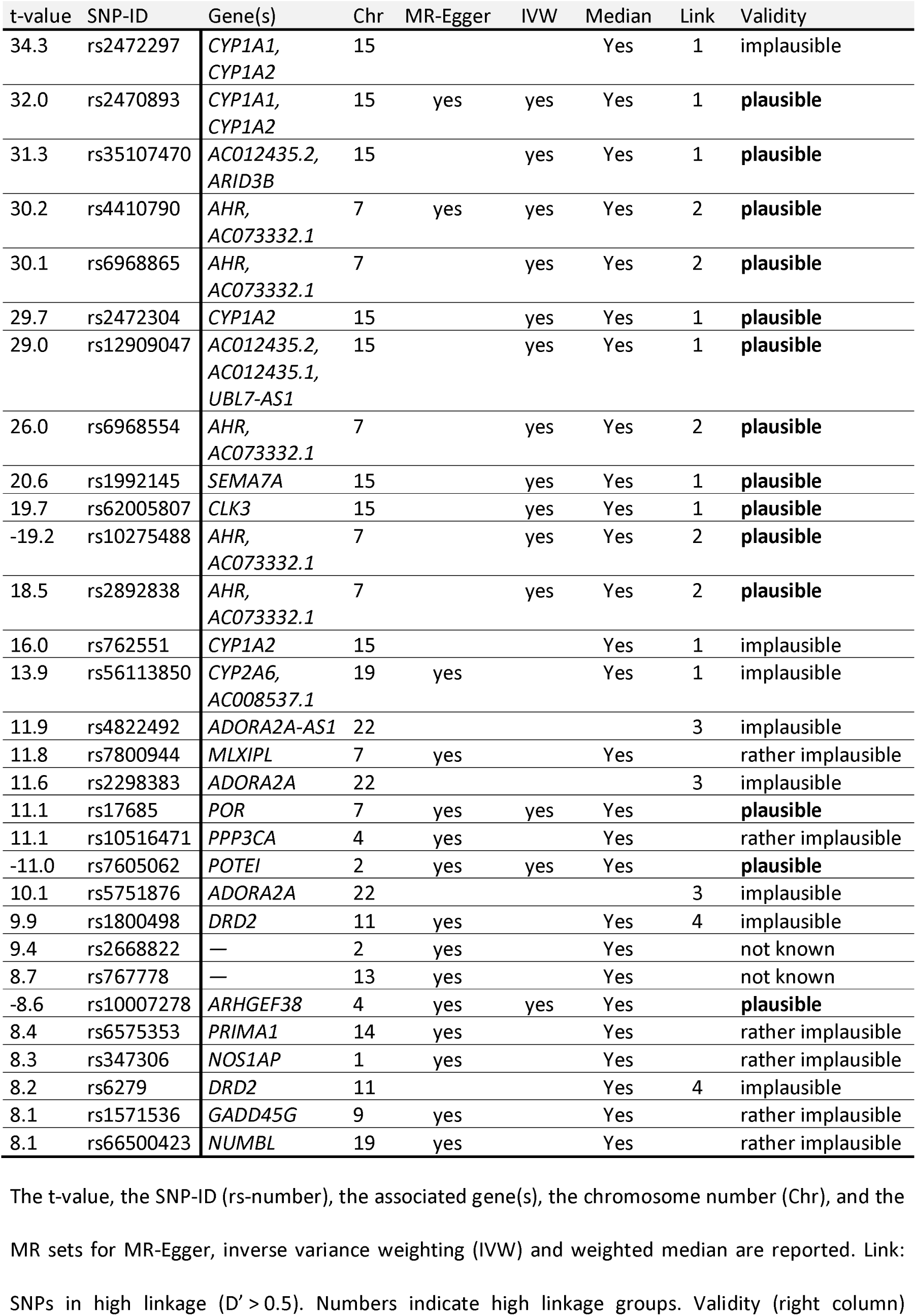
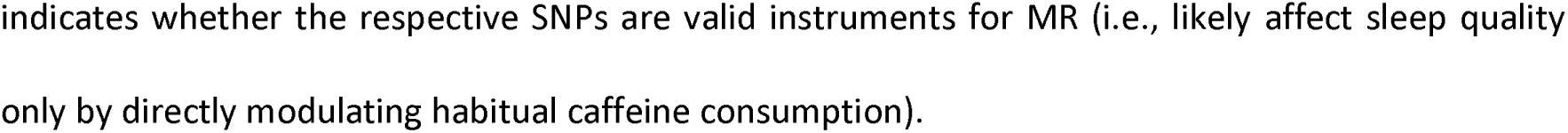
Selection of single nucleotide polymorphisms.

| t-value | SNP-ID | Gene(s) | Chr | MR-Egger | IVW | Median | Link | Validity |
| --- | --- | --- | --- | --- | --- | --- | --- | --- |
| 34.3 | rs2472297 | *CYP1A1*,<br>*CYP1A2* | 15 |  |  | Yes | 1 | implausible |
| 32.0 | rs2470893 | *CYP1A1*,<br>*CYP1A2* | 15 | yes | yes | Yes | 1 | plausible |
| 31.3 | rs35107470 | *AC012435.2*,<br>*ARID3B* | 15 |  | yes | Yes | 1 | plausible |
| 30.2 | rs4410790 | *AHR*,<br>*AC073332.1* | 7 | yes | yes | Yes | 2 | plausible |
| 30.1 | rs6968865 | *AHR*,<br>*AC073332.1* | 7 |  | yes | Yes | 2 | plausible |
| 29.7 | rs2472304 | *CYP1A2* | 15 |  | yes | Yes | 1 | plausible |
| 29.0 | rs12909047 | *AC012435.2*,<br>*AC012435.1*,<br>*UBL7-AS1* | 15 |  | yes | Yes | 1 | plausible |
| 26.0 | rs6968554 | *AHR*,<br>*AC073332.1* | 7 |  | yes | Yes | 2 | plausible |
| 20.6 | rs1992145 | *SEMA7A* | 15 |  | yes | Yes | 1 | plausible |
| 19.7 | rs62005807 | *CLK3* | 15 |  | yes | Yes | 1 | plausible |
| -19.2 | rs10275488 | *AHR*,<br>*AC073332.1* | 7 |  | yes | Yes | 2 | plausible |
| 18.5 | rs2892838 | *AHR*,<br>*AC073332.1* | 7 |  | yes | Yes | 2 | plausible |
| 16.0 | rs762551 | *CYP1A2* | 15 |  |  | Yes | 1 | implausible |
| 13.9 | rs56113850 | *CYP2A6*,<br>*AC008537.1* | 19 | yes |  | Yes | 1 | implausible |
| 11.9 | rs4822492 | *ADORA2A-AS1* | 22 |  |  |  | 3 | implausible |
| 11.8 | rs7800944 | *MLXIPL* | 7 | yes |  | Yes |  | rather implausible |
| 11.6 | rs2298383 | *ADORA2A* | 22 |  |  |  | 3 | implausible |
| 11.1 | rs17685 | *POR* | 7 | yes | yes | Yes |  | plausible |
| 11.1 | rs10516471 | *PPP3CA* | 4 | yes |  | Yes |  | rather implausible |
| -11.0 | rs7605062 | *POTEI* | 2 | yes | yes | Yes |  | plausible |
| 10.1 | rs5751876 | *ADORA2A* | 22 |  |  |  | 3 | implausible |
| 9.9 | rs1800498 | *DRD2* | 11 | yes |  | Yes | 4 | implausible |
| 9.4 | rs2668822 | — | 2 | yes |  | Yes |  | not known |
| 8.7 | rs767778 | — | 13 | yes |  | Yes |  | not known |
| -8.6 | rs10007278 | *ARHGEF38* | 4 | yes | yes | Yes |  | plausible |
| 8.4 | rs6575353 | *PRIMA1* | 14 | yes |  | Yes |  | rather implausible |
| 8.3 | rs347306 | *NOS1AP* | 1 | yes |  | Yes |  | rather implausible |
| 8.2 | rs6279 | *DRD2* | 11 |  |  | Yes | 4 | implausible |
| 8.1 | rs1571536 | *GADD45G* | 9 | yes |  | Yes |  | rather implausible |
| 8.1 | rs66500423 | *NUMBL* | 19 | yes |  | Yes |  | rather implausible |

The t-value, the SNP-ID (rs-number), the associated gene(s), the chromosome number (Chr), and the MR sets for MR-Egger, inverse variance weighting (IVW) and weighted median are reported. Link: SNPs in high linkage ( $D' > 0.5$ ). Numbers indicate high linkage groups. Validity (right column)
indicates whether the respective SNPs are valid instruments for MR (i.e., likely affect sleep quality only by directly modulating habitual caffeine consumption).

Each of the three MR methods requires different assumptions on the set of SNPs selected. To select sets likely to fulfill the assumptions, we first checked whether each of the 30 individual SNPs is plausibly a valid instrument for the causal effects of habitual caffeine consumption on objective sleep measures. Because validity cannot be verified with statistical tests, we performed a comprehensive literature search on the functions of each SNP and whether it may affect sleep directly or indirectly, i.e., not mediated by its effect on caffeine. The genes’ functionalities were either identified in papers found on Google Scholar or through the extensive gene database GeneCards^79^ and the protein database UniProt^80,81^. The associations of SNPs with sleep related topics were either identified in papers found on Google Scholar or through the NHGRI-EBI GWAS Catalog^82^ and human genetic wiki SNPedia^83^. We used this knowledgebase to construct the following validity categories: *plausible*, *rather plausible*, *unknown*, *rather implausible* and *implausible*. This categorization is not meant to be final or complete, but an attempt to screen for potentially non-valid SNPs with the help of prior biological knowledge (see supplementary Table S3 for further information).

Furthermore, certain combinations of alleles at different genetic locations can be inherited more frequently than expected by chance. This effect is called Linkage Disequilibrium (LD)^84,85^. Thus, certain SNPs can be biologically connected to each other, which affects whether they are likely to fulfill the InSIDE assumption of the MR-Egger method. One way to measure LD while retaining comparability between different pairs of alleles, is the relative LD measure D prime (D’)^86^. It measures in percentage how dependent two SNPs are. We chose D’ instead of r^2^, another measure of LD, as D’ also captures non-linear dependencies^87^ (supplementary Fig. S2). We computed D’ for each SNP pair within a given chromosome with the online tool LDmatrix from LDlink^88^ based on European populations, which included subpopulations from Great Britain and central Europe, corresponding to the locations of the cohorts used. We considered a D’ value of more than 0.5 to show high linkage as it is the halfway point of the scale.

For the MR-Egger method, we removed the variables related to adenosine receptors^35^, as these influence sleep most directly. As discussed above, MR-Egger requires a set of variables that are biologically relatively independent of each other, to fulfill the InSIDE assumption. Thus, we selected the first (based on the absolute t-value ordering) SNP member of each high LD group (D’ > 0.5), whenever there was a LD grouping present, or else single independent SNPs (Table 2 and supplementary Fig. S3). The SNP set used for IVW consisted of the plausibly valid SNPs (Table 2). For the median method, which has the least stringent requirements for SNP selection, we also removed the adenosine receptor variants due to their known direct effect on sleep (Table 2).

### Statistical analyses

We performed all analyses in R. First, we estimated the association between the SNPs and the self-reported average of caffeinated beverages per day with linear regression, adjusting for age and sex using the UK Biobank data set. We did so because age and sex are independent of the SNPs considered, but predictive of the sleep outcome variables. Adjusting for these two covariates cannot render a valid SNP invalid, but may improve the accuracy of our estimates^89,90^.

Second, we estimated the association between the SNPs and the six objective and three subjective measures of sleep quality considered, with linear regression, again adjusting for age and sex using the HypnoLaus data set. We log-transformed the outcome variables sleep latency (min), number of awakenings, the PSQI global score, and the ESS score. All these variables are strictly positive with a right-skewed distribution (Fig. 2). Furthermore, we logit-transformed NREM sigma power (%) because it displayed a heavily left-skewed distribution (Fig. 2).

We computed Wald ratio estimates for the causal effect of high habitual caffeine consumption on the six objective and three subjective measures of sleep quality introduced above (Fig. 3). To reduce the likelihood that our estimates are affected by pleiotropy, we applied three methods that combined Wald ratios to obtain a more stable estimate: MR-Egger^51^, IVW^38^, and weighted median^52^. We applied each of the three methods using selected subsets of the available SNPs (see Table 2) with the MendelianRandomization package version 0.9.0^91^ in R. We used the same method-specific SNP sets for all nine outcome variables considered.

For comparison and verification, we also estimated the causal effect of high habitual caffeine consumption on sleep quality with a causal matching estimator using the MatchingFrontier package version 4.1.0 for the matching on all available covariables and retaining the 50% best matching pairs^54^. As a baseline effect, we also computed the observational effect of habitual caffeine consumption on the nine respective outcome variables with linear regression adjusting for age and sex in the HypnoLaus data set (Fig. 3).

## Acknowledgements

We wish to thank the UK Biobank and HypnoLaus/CoLaus team for supplying us with the data and their support. The authors thank Camila Hirotsu, PhD, for her help with the data curation and analyses.

## Authors Contributions

HPL proposed and together with RH conceived the research. JHR, PMV, FS and RH collected and preprocessed the data. BS and LH processed and performed the data analyses under the supervision of MM. BS and LH created the figures and tables under the supervision of HPL. BS and LH analyzed the results and together with HPL wrote the first draft of the manuscript. All authors reviewed, edited and approved the manuscript before submission.

## Data availability

Access to both data sets is possible but requires application. For the UK Biobank application procedure see: www.ukbiobank.ac.uk/enable-your-research/apply-for-access. Information on how to access CoLaus/HypnoLaus can be found here: https://www.colaus-psycolaus.ch/professionals/how-to-collaborate.

## Code availability

The code of the analyses is available at https://github.com/henckell/HypnoLausAnalysis.

