## supplemental information for "Community-based causal evidence that high habitual caffeine consumption alters distinct polysomnography-derived sleep variables"

** indicates shared first authorship*

### Supplementary material

**Address for correspondence:**

Benjamin Stucky, PhD

Institute of Pharmacology & Toxicology

University of Zürich

Winterthurerstrasse 190

8057 Zürich, Switzerland

**Supplementary Figure S1**

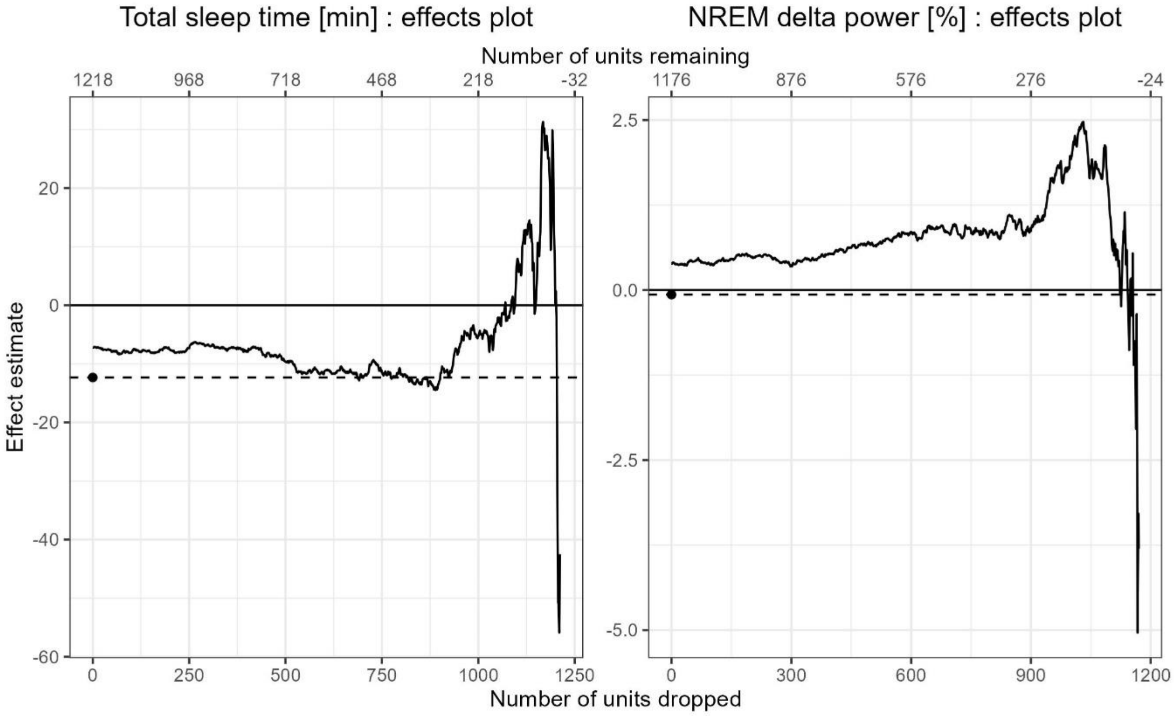


**Causal matching estimates per units dropped.** Shown are the matching estimates for the effect on total sleep time and NREM delta power as a percentage of total power per number of units dropped when using the matching from the MatchingFrontier R package.

**Supplementary Figure S2**

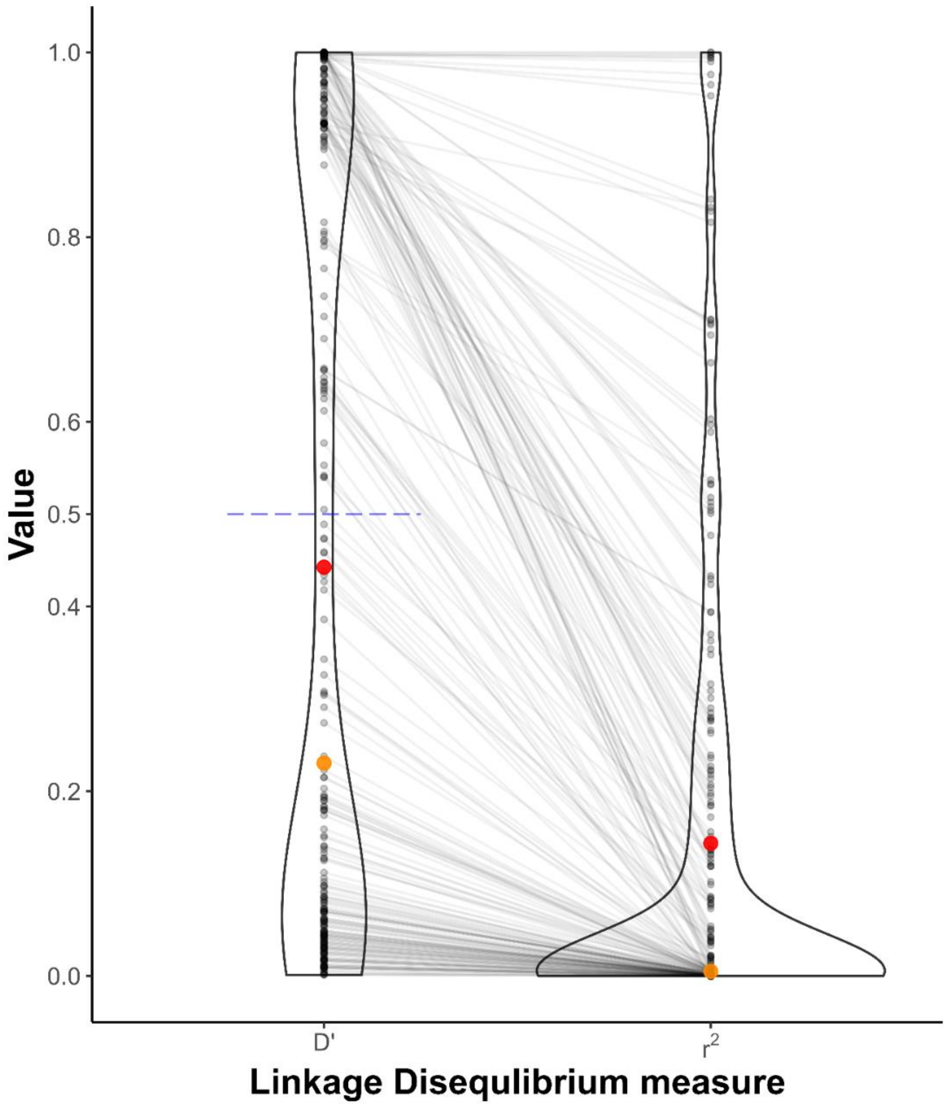


**Comparing Linkage Disequilibrium measures.** The plot shows the distribution of two relative measures of Linkage Disequilibrium, D’ and $r^2$, across all 83 selected SNPs. The red dots indicate the mean, the orange dots the median. The blue line displays the cut-off point 0.5 for D’. The grey lines between the two measures display how individual pairs of SNPs differ in their respective Linkage Disequilibrium measures. D’ exhibits a sharper separation of high to low Linkage Disequilibrium and captures non-linear linkage. The cut-off point 0.5 is the halfway point of the scale. Incidentally it is also close to the mean of the distribution.

**Supplementary Figure S3**

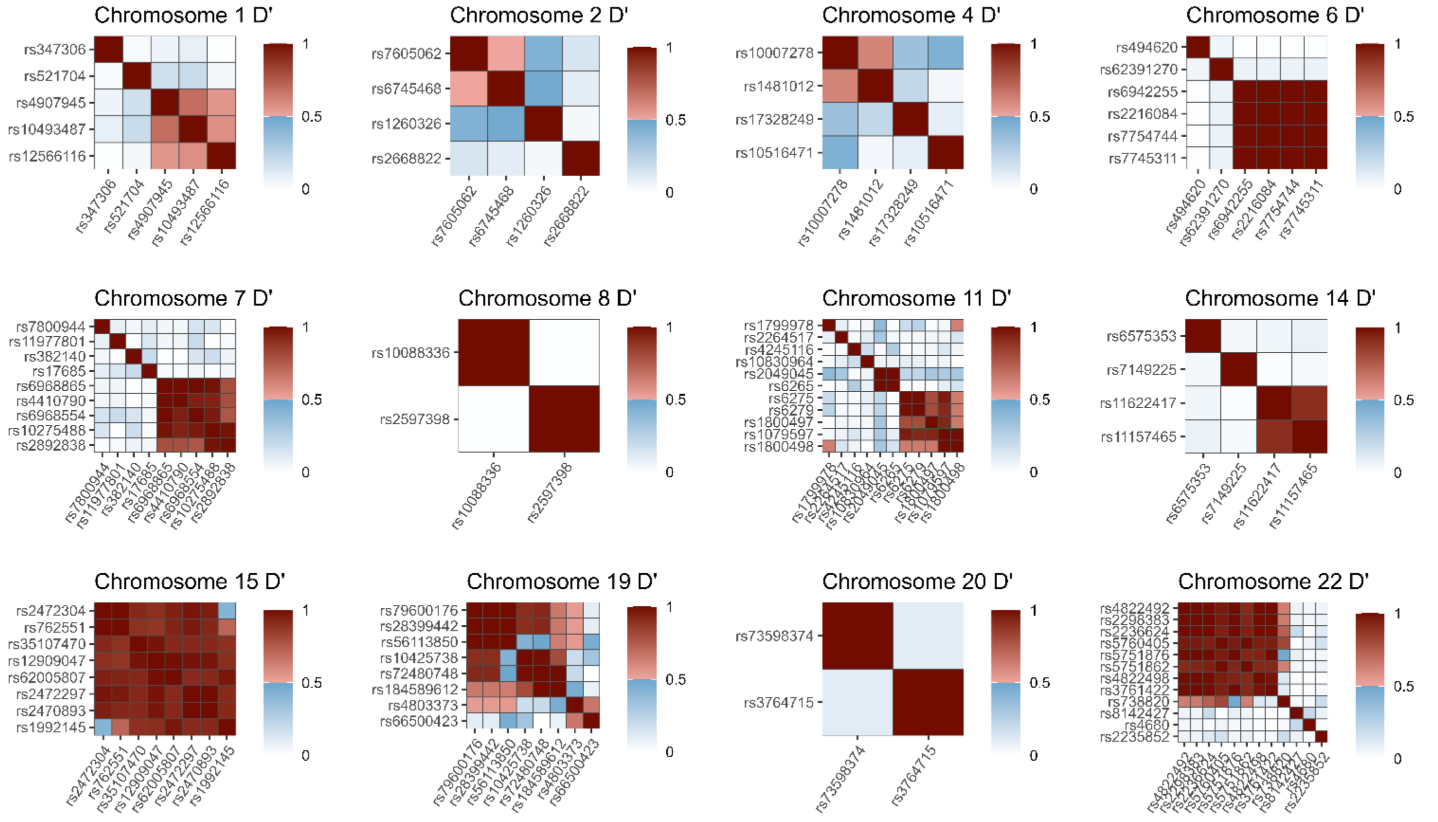


**High linkage groupings.** Shown are the D' values between each SNP in a given Chromosome from the LDlink online tool. Values below the threshold of 0.5 in blue indicate weak linkage disequilibrium and values above 0.5 in red indicate high linkage disequilibrium. Variables are grouped by hierarchical clustering to show connected groupings.

**Supplementary Table S1.** Demographic characteristics of HypnoLaus and UKBiobank samples.

| | Variable | Frequency | Overall | Moderate Intake | High Intake | High - Moderate | P (t) | P (wilcox) | P (chi-squared) |
|---|---|---|---|---|---|---|---|---|---|
| **HypnoLaus** | Sample size | | 1'726 | 1'262 | 464 | -798 | — | — | — |
| | Age [y] | | 58.54 (10.58) | 59.13 (10.63) | 56.94 (10.3) | -2.19 | <0.001 | <0.001 | — |
| | Gender | Male | 840 | 602 | 238 | 3.6 % | — | — | 0.204 |
| | | Female | 886 | 660 | 226 | -3.6 % | | | |
| | BMI | | 26.24 (4.41) | 26.23 (4.39) | 26.26 (4.48) | 0.03 | 0.902 | 0.874 | — |
| | Total sleep time [h] | | 6.67 (1.19) | 6.72 (1.18) | 6.53 (1.21) | -0.19 | 0.004 | 0.002 | — |
| | Smoking status | Non-smoker | 721 | 576 | 145 | -14.4 % | — | — | <0.001 |
| | | Current | 314 | 190 | 124 | 11.7 % | | | |
| | | Former | 688 | 494 | 194 | 2.7 % | | | |
| | Cigarettes per day | | 14.91 (10.44) | 13.39 (10.03) | 17.18 (10.67) | 3.79 | 0.003 | <0.001 | — |
| | Alcohol intake frequency | >2 / day | 28 | 21 | 7 | -0.2 % | — | — | 0.837 |
| | | 2 / day | 156 | 123 | 33 | -3.2 % | | | |
| | | 1 / day | 230 | 183 | 47 | -5.2 % | | | |
| | | 3 - 6 / week | 294 | 204 | 90 | 3.5 % | | | |
| | | 1 - 2 / week | 475 | 332 | 143 | 4.8 % | | | |
| | | Less frequent | 303 | 221 | 82 | -0.1 % | | | |
| | | Never | 6 | 3 | 3 | 0.5 % | | | |
| | Horne-Oestberg Score | | 53.23 (4.25) | 53.25 (4.21) | 53.2 (4.36) | -0.05 | 0.835 | 0.634 | — |
| | PSQI Score | | 5.08 (3.28) | 5.06 (3.25) | 5.12 (3.34) | 0.06 | 0.757 | 0.785 | — |
| | Epwort Score | | 6.11 (3.78) | 6.02 (3.71) | 6.37 (3.96) | 0.35 | 0.11 | 0.206 | — |
| **UKBiobank** | Sample size | | 485'511 | 355'914 | 129'597 | -226'317 | | | |
| | Age [y] | | 56.54 (8.09) | 56.59 (8.13) | 56.42 (7.98) | -0.17 | <0.001 | <0.001 | — |
| | Gender | Male | 222173 | 157353 | 64820 | -5.8 % | — | — | <0.001 |
| | | Female | 263338 | 198561 | 64777 | 5.8 % | | | |
| | BMI | | 27.42 (4.79) | 27.3 (4.78) | 27.74 (4.79) | 0.45 | <0.001 | <0.001 | — |
| | Total sleep time [h] | | 7.1 (1.3) | 7.12 (1.29) | 7.07 (1.24) | -0.05 | <0.001 | <0.001 | — |
| | Smoking status | Non-smoker | 264750 | 201514 | 63236 | 7.8 % | — | — | 0.721 |
| | | Current | 50960 | 31770 | 19190 | -5.9 % | | | |
| | | Former | 167969 | 121242 | 46727 | -2 % | | | |
| | | Prefer not to answer | 1817 | 1376 | 441 | 0 % | | | |
| | Cigarettes per day | | 15.37 (8.55) | 14.71 (8.49) | 16.35 (8.5) | 1.63 | <0.001 | <0.001 | — |
| | Alcohol intake frequency | Daily or almost daily | 98957 | 70687 | 28270 | -2 % | — | — | 0.368 |
| | | 3 - 4 / week | 112257 | 81316 | 30941 | -1 % | | | |
| | | 1 - 2 / week | 125272 | 92033 | 33239 | 0.2 % | | | |
| | | 1 - 3 / month | 54043 | 39542 | 14501 | -0.1 % | | | |
| | | Special occasions only | 55737 | 41888 | 13849 | 1.1 % | | | |
| | | Never | 38825 | 30138 | 8687 | 1.8 % | | | |
| | | Prefer not to answer | 405 | 298 | 107 | 0 % | | | |
| | Subjective sleeplessness | Usually | 136768 | 100528 | 36240 | 0.3 % | — | — | 0.6 |
| | | Sometimes | 231196 | 170368 | 60828 | 0.9 % | | | |
| | | Never/rarely | 117097 | 84686 | 32411 | -1.2 % | | | |
| | | Prefer not to answer | 435 | 320 | 115 | 0 % | | | |

Shown are overall values, split into the moderate (≤ 3 cups of caffeinated beverage per day) and high (≥ 4 cups of caffeinated beverage per day) caffeine intake groups, the difference (if it is a numeric value, then the mean difference, if a factor a percentual change), and the p-values for the t-test, Wilcoxon test and Chi-squared test (whenever applicable) for a variety of demographic measures in both the HypnoLaus and the UK Biobank cohorts.

**Supplementary Table S2.** Results overview.

| Variable | Panel | Method | Estimate | SE | CI 2.5% | CI 97.5% | p-value | Cohen's d |
|---|---|---|---|---|---|---|---|---|
| Log # awakenings | objective | IVW | 0.321 | 0.302 | -0.271 | 0.912 | 0.288 | |
| Log # awakenings | objective | Matching | -0.079 | 0.026 | -0.131 | -0.027 | 0.003 | -0.166 |
| Log # awakenings | objective | Median | 0.644 | 0.251 | 0.152 | 1.136 | 0.010 | |
| Log # awakenings | objective | MR Egger | 1.102 | 0.532 | 0.059 | 2.146 | 0.038 | |
| Log # awakenings | objective | Observational | -0.048 | 0.025 | -0.097 | 0.001 | 0.054 | |
| Log sleep latency [min] | objective | IVW | 0.965 | 0.576 | -0.163 | 2.094 | 0.094 | |
| Log sleep latency [min] | objective | Matching | -0.132 | 0.057 | -0.244 | -0.021 | 0.020 | -0.136 |
| Log sleep latency [min] | objective | Median | 0.461 | 0.488 | -0.496 | 1.417 | 0.345 | |
| Log sleep latency [min] | objective | MR Egger | 1.109 | 1.212 | -1.267 | 3.484 | 0.360 | |
| Log sleep latency [min] | objective | Observational | -0.056 | 0.048 | -0.151 | 0.038 | 0.244 | |
| Logit NREM sigma [%] | objective | IVW | 0.166 | 0.529 | -0.870 | 1.202 | 0.753 | |
| Logit NREM sigma [%] | objective | Matching | -0.024 | 0.031 | -0.086 | 0.037 | 0.440 | -0.046 |
| Logit NREM sigma [%] | objective | Median | 0.146 | 0.350 | -0.541 | 0.832 | 0.678 | |
| Logit NREM sigma [%] | objective | MR Egger | 0.561 | 0.882 | -1.168 | 2.291 | 0.525 | |
| Logit NREM sigma [%] | objective | Observational | -0.029 | 0.031 | -0.089 | 0.031 | 0.348 | |
| Logit sleep efficiency [%] | objective | IVW | -0.385 | 0.493 | -1.351 | 0.580 | 0.434 | |
| Logit sleep efficiency [%] | objective | Matching | 0.058 | 0.044 | -0.029 | 0.144 | 0.193 | 0.068 |
| Logit sleep efficiency [%] | objective | Median | -0.216 | 0.417 | -1.033 | 0.602 | 0.605 | |
| Logit sleep efficiency [%] | objective | MR Egger | -0.727 | 0.879 | -2.449 | 0.996 | 0.408 | |
| Logit sleep efficiency [%] | objective | Observational | 0.048 | 0.041 | -0.032 | 0.128 | 0.238 | |
| NREM delta [%] | objective | IVW | 8.815 | 3.881 | 1.209 | 16.421 | 0.023 | |
| NREM delta [%] | objective | Matching | 0.827 | 0.348 | 0.144 | 1.510 | 0.018 | 0.140 |
| NREM delta [%] | objective | Median | 10.250 | 3.265 | 3.850 | 16.649 | 0.002 | |
| NREM delta [%] | objective | MR Egger | 3.172 | 6.900 | -10.351 | 16.695 | 0.646 | |
| NREM delta [%] | objective | Observational | 0.019 | 0.329 | -0.627 | 0.665 | 0.954 | |
| REM sleep [%] | objective | IVW | -3.820 | 3.902 | -11.469 | 3.828 | 0.328 | |
| REM sleep [%] | objective | Matching | -0.150 | 0.331 | -0.800 | 0.500 | 0.651 | -0.025 |
| REM sleep [%] | objective | Median | 3.586 | 3.541 | -3.354 | 10.527 | 0.311 | |
| REM sleep [%] | objective | MR Egger | -0.503 | 7.115 | -14.449 | 13.443 | 0.944 | |
| REM sleep [%] | objective | Observational | -0.369 | 0.321 | -0.998 | 0.260 | 0.250 | |
| Total sleep time [min] | objective | IVW | -124.980 | 45.077 | -213.330 | -36.631 | 0.006 | |
| Total sleep time [min] | objective | Matching | -10.980 | 4.070 | -18.973 | -2.987 | 0.007 | -0.155 |
| Total sleep time [min] | objective | Median | -139.572 | 37.602 | -213.270 | -65.873 | 0.000 | |
| Total sleep time [min] | objective | MR Egger | -229.358 | 80.860 | -387.841 | -70.875 | 0.005 | |
| Total sleep time [min] | objective | Observational | -12.904 | 3.760 | -20.279 | -5.529 | 0.001 | |
| Log ESS score | subjective | IVW | -0.850 | 0.415 | -1.663 | -0.036 | 0.041 | |
| Log ESS score | subjective | Matching | 0.097 | 0.025 | 0.047 | 0.147 | 0.000 | 0.148 |
| Log ESS score | subjective | Median | -0.468 | 0.339 | -1.133 | 0.197 | 0.167 | |
| Log ESS score | subjective | MR Egger | -2.159 | 1.137 | -4.387 | 0.069 | 0.057 | |
| Log ESS score | subjective | Observational | 0.018 | 0.035 | -0.050 | 0.086 | 0.595 | |
| Log PSQI global score | subjective | IVW | -0.193 | 0.391 | -0.958 | 0.573 | 0.622 | |
| Log PSQI global score | subjective | Matching | 0.052 | 0.026 | 0.000 | 0.104 | 0.051 | 0.077 |
| Log PSQI global score | subjective | Median | -0.314 | 0.305 | -0.912 | 0.284 | 0.304 | |
| Log PSQI global score | subjective | MR Egger | -0.307 | 0.684 | -1.647 | 1.032 | 0.653 | |
| Log PSQI global score | subjective | Observational | 0.018 | 0.032 | -0.044 | 0.080 | 0.564 | |
| MEQ score | subjective | IVW | 3.355 | 3.040 | -2.604 | 9.314 | 0.270 | |
| MEQ score | subjective | Matching | 0.054 | 0.166 | -0.272 | 0.381 | 0.743 | 0.013 |
| MEQ score | subjective | Median | 8.338 | 2.817 | 2.816 | 13.860 | 0.003 | |
| MEQ score | subjective | MR Egger | 10.183 | 7.343 | -4.210 | 24.576 | 0.166 | |
| MEQ score | subjective | Observational | 0.030 | 0.246 | -0.452 | 0.513 | 0.901 | |

Shown are the results for the Mendelian Randomization methods MR-Egger, Inverse variance weighting (IVW) and Median, the causal matching estimator (MatchingFrontier) and the observational linear regression estimator. The objective and subjective variable panels correspond to those in the main paper. Outcome variables include, total sleep time, log sleep latency in minutes, delta power in NREM, logit sigma power in NREM and REM sleep in percentages, log number of awakenings, log ESS score, log PSQI global score and the MEQ score. We provide the effect estimates with their standard errors (SE), the 95% confidence interval, the corresponding p-value and the Cohen's d effect size for the matching estimator.

**Supplementary Table S3.** Summary of pre-selected single nucleotide polymorphisms.

| t | SNP-id | Gene(s) | Chr | IVW | Median | MR Egger | Linkage | Validity | Interactions | Publications on Implausibility |
|---|---|---|---|---|---|---|---|---|---|---|
| 34.3 | rs2472297 | *CYP1A1, CYP1A2* | 15 | | yes | | 1 | implausible | Alcohol | (Zhou et al., 2020)<br>(Liu et al., 2019)<br>(Zhong et al., 2019) |
| 32.0 | rs2470893 | *CYP1A1, CYP1A2* | 15 | yes | yes | yes | 1 | plausible | | |
| 31.3 | rs35107470 | *AC012435.2, ARID3B* | 15 | yes | yes | | 1 | plausible | | |
| 30.2 | rs4410790 | *AHR, AC073332.1* | 7 | yes | yes | yes | 2 | plausible | | |
| 30.1 | rs6968865 | *AHR, AC073332.1* | 7 | yes | yes | | 2 | plausible | | |
| 29.7 | rs2472304 | *CYP1A2* | 15 | yes | yes | | 1 | plausible | | |
| 29.0 | rs12909047 | *AC012435.2, AC012435.1, UBL7-AS1* | 15 | yes | yes | | 1 | plausible | | |
| 26.0 | rs6968554 | *AHR, AC073332.1* | 7 | yes | yes | | 2 | plausible | | |
| 20.6 | rs1992145 | *SEMA7A* | 15 | yes | yes | | 1 | plausible | | |
| 19.7 | rs62005807 | *CLK3* | 15 | yes | yes | | 1 | plausible | | |
| -19.2 | rs10275488 | *AHR, AC073332.1* | 7 | yes | yes | | 2 | plausible | | |
| 18.5 | rs2892838 | *AHR, AC073332.1* | 7 | yes | yes | | 2 | plausible | | |
| 16.0 | rs762551 | *CYP1A2* | 15 | | yes | | 1 | implausible | Smoking | (Wang et al., 2013)<br>(Sachse et al., 1999) |
| 13.9 | rs56113850 | *CYP2A6, AC008537.1* | 19 | | yes | yes | 3 | implausible | Smoking | (Buchwald et al., 2021)<br>(Loukola et al., 2015)<br>(Patel et al., 2016)<br>(McKay et al., 2017) |
| 11.9 | rs4822492 | *ADORA2A-AS1* | 22 | | | | 4 | implausible | Adenosine | (Erblang et al., 2019) |
| 11.8 | rs7800944 | *MLXIPL* | 7 | | yes | yes | | rather implausible | Williams Beuren syndrome | (Goldman et al., 2009) |
| 11.6 | rs2298383 | *ADORA2A* | 22 | | | | 4 | implausible | Adenosine | (Erblang et al., 2019) |
| 11.1 | rs17685 | *POR* | 7 | yes | yes | yes | | plausible | | |
| 11.1 | rs10516471 | *PPP3CA* | 4 | | yes | yes | | rather implausible | Diabetes | (Meigs et al., 2007) |
| -11.0 | rs7605062 | *POTEI* | 2 | yes | yes | yes | 5 | plausible | | |
| 10.1 | rs5751876 | *ADORA2A* | 22 | | | | 4 | implausible | Adenosine | (Erblang et al., 2019) |
| 9.9 | rs1800498 | *DRD2* | 11 | | yes | yes | 6 | implausible | Dopamine | (Monti and Monti, 2007) |
| 9.4 | rs2668822 | — | 2 | | yes | yes | | unknown | | |
| 8.7 | rs767778 | — | 13 | | yes | yes | | unknown | | |
| -8.6 | rs10007278 | *ARHGEF38* | 4 | yes | yes | yes | 7 | plausible | | |
| 8.4 | rs6575353 | *PRIMA1* | 14 | | yes | yes | | rather implausible | Acetylcholin | (Hildebrand et al., 2015)<br>(Watson et al., 2010) |
| 8.3 | rs347306 | *NOS1AP* | 1 | | yes | yes | | rather implausible | Depression, Schizophrenia | (Cheah et al., 2015) |
| 8.2 | rs6279 | *DRD2* | 11 | | yes | | 6 | implausible | Dopamine, Alcohol | (Monti and Monti, 2007)<br>(Meyers et al., 2013) |
| 8.1 | rs1571536 | *GADD45G* | 9 | | yes | yes | | rather implausible | Cellular stress and sleep | (Naidoo, 2009) |
| 8.1 | rs66500423 | *NUMBL* | 19 | | yes | yes | 8 | rather implausible | Nicotine Dependence | (Hatoum et al., 2023) |
| 8.0 | rs6275 | *DRD2* | 11 | | | | 6 | | | |
| -7.8 | rs6745468 | *EMX1* | 2 | | | | 5 | | | |
| -7.4 | rs2270969 | *MCCC1* | 3 | | | | | | | |

| | | | | |
|---|---|---|---|---|
| -6.9 | rs7745311 | *PDSS2* | 6 | 9 |
| -6.9 | rs9386630 | *PDSS2* | 6 | 9 |
| -6.8 | rs7754744 | *PDSS2, RPS24P12* | 6 | 9 |
| -6.8 | rs6942255 | *PDSS2* | 6 | 9 |
| -6.8 | rs2216084 | *PDSS2* | 6 | 9 |
| 6.5 | rs5751862 | *SPECC1L* | 22 | 4 |
| 6.4 | rs17328249 | *HAND2-AS1* | 4 | |
| -6.4 | rs494620 | *SLC44A4* | 6 | |
| 6.3 | rs521704 | *near GBP4* | 1 | |
| 5.5 | rs62391270 | *RNU7-133P, AL353152.1* | 6 | |
| -5.4 | rs9902453 | *EFCAB5* | 17 | |
| -5.2 | rs1481012 | *ABCG2* | 4 | 7 |
| -5.1 | rs2264517 | — | 11 | |
| -4.6 | rs1799978 | *DRD2* | 11 | 6 |
| -4.6 | rs1800497 | *ANKK1* | 11 | 6 |
| 4.3 | rs4822498 | *ADORA2A-AS1* | 22 | 4 |
| -4.2 | rs12566116 | — | 1 | 10 |
| 4.1 | rs382140 | *NRCAM, LAMB4* | 7 | |
| 3.6 | rs72480748 | *CYP2A7P2* | 19 | 3 |
| 3.5 | rs4803373 | *CYP2F2P, AC008537.1* | 19 | 8 |
| 3.4 | rs2049045 | *BDNF, BDNF-AS* | 11 | 11 |
| 3.1 | rs2597979 | *PRH1, TAS2R14, AC018630.2* | 12 | |
| 2.6 | rs5760405 | *SPECC1L* | 22 | 4 |
| -2.6 | rs7149225 | *NRXN3* | 14 | |
| -2.6 | rs18458961 2 | *CYP2A7P2, CYP2G1P* | 19 | 3 |
| 2.3 | rs10425738 | *CYP2A7P2, CYP2B7P* | 19 | 3 |
| 1.9 | rs3761422 | *ADORA2A* | 22 | 4 |
| -1.9 | rs1079597 | *DRD2* | 11 | 6 |
| -1.8 | rs4239278 | — | 18 | |
| -1.6 | rs2236624 | *ADORA2A* | 22 | 4 |
| 1.6 | rs11977801 | — | 7 | |
| 1.4 | rs4907945 | — | 1 | 10 |
| 1.4 | rs1260326 | *GCKR* | 2 | |
| -1.3 | rs28399442 | *CYP2A6, AC008537.1* | 19 | 3 |
| -1.3 | rs79600176 | *CYP2A7, AC008537.1* | 19 | 3 |
| -1.2 | rs10493487 | *NEGR1* | 1 | 10 |
| -1.2 | rs6265 | *BDNF, BDNF-AS* | 11 | 11 |

| | | | | |
|---|---|---|---|---|
| -0.9 | rs73598374 | *ADA* | 20 | |
| -0.9 | rs11157465 | — | 14 | 12 |
| -0.8 | rs2597398 | *LOC105379315* | 8 | |
| 0.8 | rs11622417 | *LINC00871* | 14 | 12 |
| 0.7 | rs10830964 | *MTNR1B, RPL26P31* | 11 | |
| 0.6 | rs10088336 | — | 8 | |
| -0.2 | rs4245116 | *OPCML* | 11 | |
| — | rs11863088 | *ATF7IP2* | 16 | |
| — | rs2235852 | *RANGAP1* | 22 | |
| — | rs3764715 | *SNPH* | 20 | |
| — | rs4680 | *COMT* | 22 | |
| — | rs738820 | *GUCD1* | 22 | 4 |
| — | rs8142427 | *FAM19A5* | 22 | |

The table provides an overview of the 83 preselected SNPs and our decision regarding which to use for each MR method. The first row contains the t-values of the linear SNP-treatment regression sorted by absolute value. The high-linkage groups (D’ > 0.5) are color coded in the linkage column. Relevant publications regarding the implausibility of a SNPs validity are included for the first 30 SNPs, i.e., those SNPs that fulfill the inclusion criteria of having an absolute t-value larger than 8.

Liu, M., Jiang, Y., Wedow, R., Li, Y., Brazel, D.M., Chen, F., Datta, G., Davila-Velderrain, J., McGuire, D., Tian, C., Zhan, X., Choquet, H., Docherty, A.R., Faul, J.D., Foerster, J.R., Fritsche, L.G., Gabrielsen, M.E., Gordon, S.D., Haessler, J., Hottenga, J.-J., Huang, H., Jang, S.-K., Jansen, P.R., Ling, Y., Mägi, R., Matoba, N., McMahon, G., Mulas, A., Orrù, V., Palviainen, T., Pandit, A., Reginsson, G.W., Skogholt, A.H., Smith, J.A., Taylor, A.E., Turman, C., Willemsen, G., Young, H., Young, K.A., Zajac, G.J.M., Zhao, W., Zhou, W., Bjornsdottir, G., Boardman, J.D., Boehnke, M., Boomsma, D.I., Chen, C., Cucca, F., Davies, G.E., Eaton, C.B., Ehringer, M.A., Esko, T., Fiorillo, E., Gillespie, N.A., Gudbjartsson, D.F., Haller, T., Harris, K.M., Heath, A.C., Hewitt, J.K., Hickie, I.B., Hokanson, J.E., Hopfer, C.J., Hunter, D.J., Iacono, W.G., Johnson, E.O., Kamatani, Y., Kardia, S.L.R., Keller, M.C., Kellis, M., Kooperberg, C., Kraft, P., Krauter, K.S., Laakso, M., Lind, P.A., Loukola, A., Lutz, S.M., Madden, P.A.F., Martin, N.G., McGue, M., McQueen, M.B., Medland, S.E., Metspalu, A., Mohlke, K.L., Nielsen, J.B., Okada, Y., Peters, U., Polderman, T.J.C., Posthuma, D., Reiner, A.P., Rice, J.P., Rimm, E., Rose, R.J., Runarsdottir, V., Stallings, M.C., Stančáková, A., Stefansson, H., Thai, K.K., Tindle, H.A., Tyrfingsson, T., Wall, T.L., Weir, D.R., Weisner, C., Whitfield, J.B., Winsvold, B.S., Yin, J., Zuccolo, L., Bierut, L.J., Hveem, K., Lee, J.J., Munafò, M.R., Saccone, N.L., Willer, C.J., Cornelis, M.C., David, S.P., Hinds, D.A., Jorgenson, E., Kaprio, J., Stitzel, J.A., Stefansson, K., Thorgeirsson, T.E., Abecasis, G., Liu, D.J., Vrieze, S., 2019. Association studies of up to 1.2 million individuals yield new insights into the genetic etiology of tobacco and alcohol use. Nat. Genet. 51, 237–244. https://doi.org/10.1038/s41588-018-0307-5

Loukola, A., Buchwald, J., Gupta, R., Palviainen, T., Hällfors, J., Tikkanen, E., Korhonen, T., Ollikainen, M., Sarin, A.-P., Ripatti, S., Lehtimäki, T., Raitakari, O., Salomaa, V., Rose, R.J., Tyndale, R.F., Kaprio, J., 2015. A Genome-Wide Association Study of a Biomarker of Nicotine Metabolism. PLOS Genet. 11, e1005498. https://doi.org/10.1371/journal.pgen.1005498

McKay, J.D., Hung, R.J., Han, Y., Zong, X., Carreras-Torres, R., Christiani, D.C., Caporaso, N.E., Johansson, Mattias, Xiao, X., Li, Y., Byun, J., Dunning, A., Pooley, K.A., Qian, D.C., Ji, X., Liu, G., Timofeeva, M.N., Bojesen, S.E., Wu, X., Le Marchand, L., Albanes, D., Bickeböller, H., Aldrich, M.C., Bush, W.S., Tardon, A., Rennert, G., Teare, M.D., Field, J.K., Kiemeney, L.A., Lazarus, P., Haugen, A., Lam, S., Schabath, M.B., Andrew, A.S., Shen, H., Hong, Y.-C., Yuan, J.-M., Bertazzi, P.A., Pesatori, A.C., Ye, Y., Diao, N., Su, L., Zhang, R., Brhane, Y., Leighl, N., Johansen, J.S., Mellemgaard, A., Saliba, W., Haiman, C.A., Wilkens, L.R., Fernandez-Somoano, A., Fernandez-Tardon, G., van der Heijden, H.F.M., Kim, J.H., Dai, J., Hu, Z., Davies, M.P.A., Marcus, M.W., Brunnström, H., Manjer, J., Melander, O., Muller, D.C., Overvad, K., Trichopoulou, A., Tumino, R., Doherty, J.A., Barnett, M.P., Chen, C., Goodman, G.E., Cox, A., Taylor, F., Woll, P., Brüske, I., Wichmann, H.-E., Manz, J., Muley, T.R., Risch, A., Rosenberger, A., Grankvist, K., Johansson, Mikael, Shepherd, F.A., Tsao, M.-S., Arnold, S.M., Haura, E.B., Bolca, C., Holcatova, I., Janout, V., Kontic, M., Lissowska, J., Mukeria, A., Ognjanovic, S., Orlowski, T.M., Scelo, G., Swiatkowska, B., Zaridze, D., Bakke, P., Skaug, V., Zienolddiny, S., Duell, E.J., Butler, L.M., Koh, W.-P., Gao, Y.-T., Houlston, R.S., McLaughlin, J., Stevens, V.L., Joubert, P., Lamontagne, M., Nickle, D.C., Obeidat, M., Timens, W., Zhu, B., Song, L., Kachuri, L., Artigas, M.S., Tobin, M.D., Wain, L.V., Rafnar, T., Thorgeirsson, T.E., Reginsson, G.W., Stefansson, K., Hancock, D.B., Bierut, L.J., Spitz, M.R., Gaddis, N.C., Lutz, S.M., Gu, F., Johnson, E.O., Kamal, A., Pikielny, C., Zhu, D., Lindströem, S., Jiang, X., Tyndale, R.F., Chenevix-Trench, G., Beesley, J., Bossé, Y., Chanock, S., Brennan, P., Landi, M.T., Amos, C.I., 2017. Large-scale association analysis identifies new lung cancer susceptibility loci and heterogeneity in genetic susceptibility across histological subtypes. Nat. Genet. 49, 1126–1132. https://doi.org/10.1038/ng.3892

Meigs, J.B., Manning, A.K., Fox, C.S., Florez, J.C., Liu, C., Cupples, L.A., Dupuis, J., 2007. Genome-wide association with diabetes-related traits in the Framingham Heart Study. BMC Med. Genet. 8, 1–10. https://doi.org/10.1186/1471-2350-8-S1-S16

Meyers, J.L., Nyman, E., Loukola, A., Rose, R.J., Kaprio, J., Dick, D.M., 2013. The association between DRD2/ANKK1 and genetically informed measures of alcohol use and problems. Addict. Biol. 18, 523–536. https://doi.org/10.1111/j.1369-1600.2012.00490.x

Monti, J.M., Monti, D., 2007. The involvement of dopamine in the modulation of sleep and waking. Sleep Med. Rev. 11, 113–133. https://doi.org/10.1016/j.smrv.2006.08.003

Naidoo, N., 2009. Cellular stress/the unfolded protein response: Relevance to sleep and sleep disorders. Sleep Med. Rev. 13, 195–204. https://doi.org/10.1016/j.smrv.2009.01.001

Patel, Y.M., Park, S.L., Han, Y., Wilkens, L.R., Bickeböller, H., Rosenberger, A., Caporaso, N., Landi, M.T., Brüske, I., Risch, A., Wei, Y., Christiani, D.C., Brennan, P., Houlston, R., McKay, J., McLaughlin, J., Hung, R., Murphy, S., Stram, D.O., Amos, C., Le Marchand, L., 2016. Novel Association of Genetic Markers Affecting CYP2A6 Activity and Lung Cancer Risk. Cancer Res. 76, 5768–5776. https://doi.org/10.1158/0008-5472.CAN-16-0446

Sachse, C., Brockmöller, J., Bauer, S., Roots, I., 1999. Functional significance of a C→A polymorphism in intron 1 of the cytochrome P450 CYP1A2 gene tested with caffeine. Br. J. Clin. Pharmacol. 47, 445–449. https://doi.org/10.1046/j.1365-2125.1999.00898.x

Wang, L., Hu, Z., Deng, X., Wang, Y., Zhang, Z., Cheng, Z.-N., 2013. Association between Common CYP1A2 Polymorphisms and Theophylline Metabolism in Non-smoking Healthy Volunteers. Basic Clin. Pharmacol. Toxicol. 112, 257–263. https://doi.org/10.1111/bcpt.12038

Watson, C.J., Baghdoyan, H.A., Lydic, R., 2010. Neuropharmacology of Sleep and Wakefulness. Sleep Med. Clin. 5, 513–528. https://doi.org/10.1016/j.jsmc.2010.08.003

Zhong, V.W., Kuang, A., Danning, R.D., Kraft, P., van Dam, R.M., Chasman, D.I., Cornelis, M.C., 2019. A genome-wide association study of bitter and sweet beverage consumption. Hum. Mol. Genet. 28, 2449–2457. https://doi.org/10.1093/hmg/ddz061

Zhou, H., Sealock, J.M., Sanchez-Roige, S., Clarke, T.-K., Levey, D.F., Cheng, Z., Li, B., Polimanti, R., Kember, R.L., Smith, R.V., Thygesen, J.H., Morgan, M.Y., Atkinson, S.R., Thursz, M.R., Nyegaard, M., Mattheisen, M., Børglum, A.D., Johnson, E.C., Justice, A.C., Palmer, A.A., McQuillin, A., Davis, L.K., Edenberg, H.J., Agrawal, A., Kranzler, H.R., Gelernter, J., 2020. Genome-wide meta-analysis of problematic alcohol use in 435,563 individuals yields insights into biology and relationships with other traits. Nat. Neurosci. 23, 809–818. https://doi.org/10.1038/s41593-020-0643-5
